# Clinical outcomes of SARS-CoV-2 pandemic in long-term care facilities for people with epilepsy: observational study

**DOI:** 10.1101/2020.06.10.20123281

**Authors:** Simona Balestrini, Matthias J Koepp, Sonia Gandhi, Hannah M Rickman, Gee Yen Shin, Catherine F Houlihan, Jonny Anders-Cannon, Katri Silvennoinen, Fenglai Xiao, Sara Zagaglia, Kirsty Hudgell, Mariusz Ziomek, Paul Haimes, Adam Sampson, Annie Parker, for The Meath, Eleni Nastouli, Charles Swanton, for the CCC, Josemir W Sander, Sanjay M Sisodiya, for the ASAP Consortium

## Abstract

**Objectives:** COVID-19 is spreading in long-term care facilities with devastating outcomes worldwide, especially for people with chronic health conditions. There is a pressing need to adopt effective measures prevention and containment of in such settings.

**Design:** Retrospective cohort study assessing the effect of enhanced surveillance and early preventative strategies and comparing outcomes for people with severe epilepsy and other comorbidities

**Setting:** Three long-term care facilities: Chalfont Centre for Epilepsy (CCE), St. Elisabeth (STE), and The Meath (TM) with different models of primary and specialist care involvement, in the United Kingdom

**Participants:** 286 long-term residents (age range 19–91 years), 740 carers who had been in contact with the residents during the observation period between 16 March and 05 June 2020.

**Interventions:** Early preventative and infection control measures with identification and isolation of symptomatic cases, with additional enhanced surveillance and isolation of asymptomatic residents and carers at one site (CCE)

**Main outcome measures:** Infection rate for SARS-CoV-2 among residents and carers, asymptomatic rate and case fatality rate, if available.

**Results:** During a 12-week observation period, we identified 29 people (13 residents) who were SARS-CoV-2 positive with confirmed outbreaks amongst residents in two long-term care facilities (CCE, STE). At CCE, two out of 98 residents were symptomatic and tested positive, one of whom died. A further seven individuals testing positive on weekly enhanced surveillance had a completely asymptomatic course. One asymptomatic carer tested positive after contact with confirmed COVID-19 patients in another institution. Since 30 April 2020, during on-site weekly enhanced surveillance all 275 caregivers tested repeatedly negative. At STE, three out of 146 residents were symptomatic and tested positive, a fourth tested positive during hospital admission for symptoms not related to COVID-19. Since April 6, 2020, 105/215 carers presenting with typical symptoms for COVID-19 were tested, of whom 15 tested positive. At TM, testing of symptomatic carers only started from early/mid-April, whilst on-site testing, even of symptomatic residents, was not available until recently.

During the observation period, eight of 80 residents were symptomatic but none was tested. Twenty-six of 250 carers were symptomatic and were tested, of whom two tested positive.

**Conclusions:** Infection outbreaks in long-term care facilities for vulnerable people with epilepsy can be quickly contained, but only if asymptomatic cases are identified through enhanced surveillance at individual and care staff level. We observed a low rate of morbidity and mortality which confirmed that preventative measures with isolation of suspected and confirmed cases of COVID-19 can reduce resident-to-resident and reverse resident-to-carer transmission.

## Introduction

Novel coronavirus disease 2019 (COVID-19) associated with the SARS-CoV-2 virus has quickly spread around the world.^1^ A range of typical symptoms is associated with COVID-19, including fever, cough, and dyspnoea,^2^ but these may be absent in older age, and in those with multi-morbidity.^3^ Long-term care facilities are high-risk settings for poor outcomes from respiratory disease outbreaks, including COVID-19, due to greater prevalence of risk factors, like age and chronic health conditions.^4–6^

Until recently, only people admitted to hospital were tested for COVID-19 in the United Kingdom (UK). Frail people and those with multi-morbidity living in care-facilities were not tested, often dying in care setting. Official figures for the number of deaths in the community do not provide a comprehensive account of what has happened in care-facilities.^7^ These figures are likely to be underestimations due to the lack of testing.

Once COVID-19 is introduced into a care-facility, it has the potential to spread rapidly and widely, causing serious adverse outcomes among those in care and those providing it.^8–10^ Shielding of vulnerable people is difficult, and self-isolation, if symptomatic, almost impossible because of limited capacity. There is a higher threshold for frail people and those with challenging behaviour in long-term care to be admitted to hospital. These individuals often have restricted access to intensive care, resulting in infectious individuals remaining in communal living facilities for longer.

Asymptomatic transmission of SARS-CoV-2 is considered the Achilles’ heel of the COVID-19 pandemic.^11^ Typical care facility practices potentially contribute to greater spread, such as dependency on temporary workers who move between sites, or frequent redistribution and high turnover levels of caregivers within the same facility. Along with widespread lack of personal protection equipment (PPE), general tasks, like medication dispensation, are often performed by a few trained workers, who, if infected, may spread the infection.

Here, we report the effect of early preventative measures and enhanced surveillance in a long-term care facility for people with epilepsy and multiple co-morbidities, and compare infection rates and outcomes with three other such facilities, who all adopted similar preventative measures, including attempts at shielding vulnerable and isolating symptomatic people, but did not have access to enhanced surveillance, and only very limited access to testing even symptomatic people.

## Methods

### Approval

This work was registered and independently approved at UCLH by the Queen Square Quality & Safety Committee as a service evaluation. This approval waives the need for approval by an IRB/ethics committee, in accordance with UK legislation and NHS operating procedures.

### Sites

The Chalfont Centre for Epilepsy (CCE), north-west of London, is a long-term care facility for adults with severe epilepsy and other comorbidities. It currently houses 98 people (66 males) aged between 23 – 91 (median age: 49 years), who live in seven units of 1–4 self-contained flats, each housing 5–12 people, looked after by 275 carers during the observation period. University College London Hospitals (UCLH) provides secondary and tertiary care to people living at the centre, which also houses a UCLH elective unit for multidisciplinary assessment and treatment of adults with complex epilepsies (Sir William Gowers Centre, SWGC) (Figure 1).

**Figure 1.**
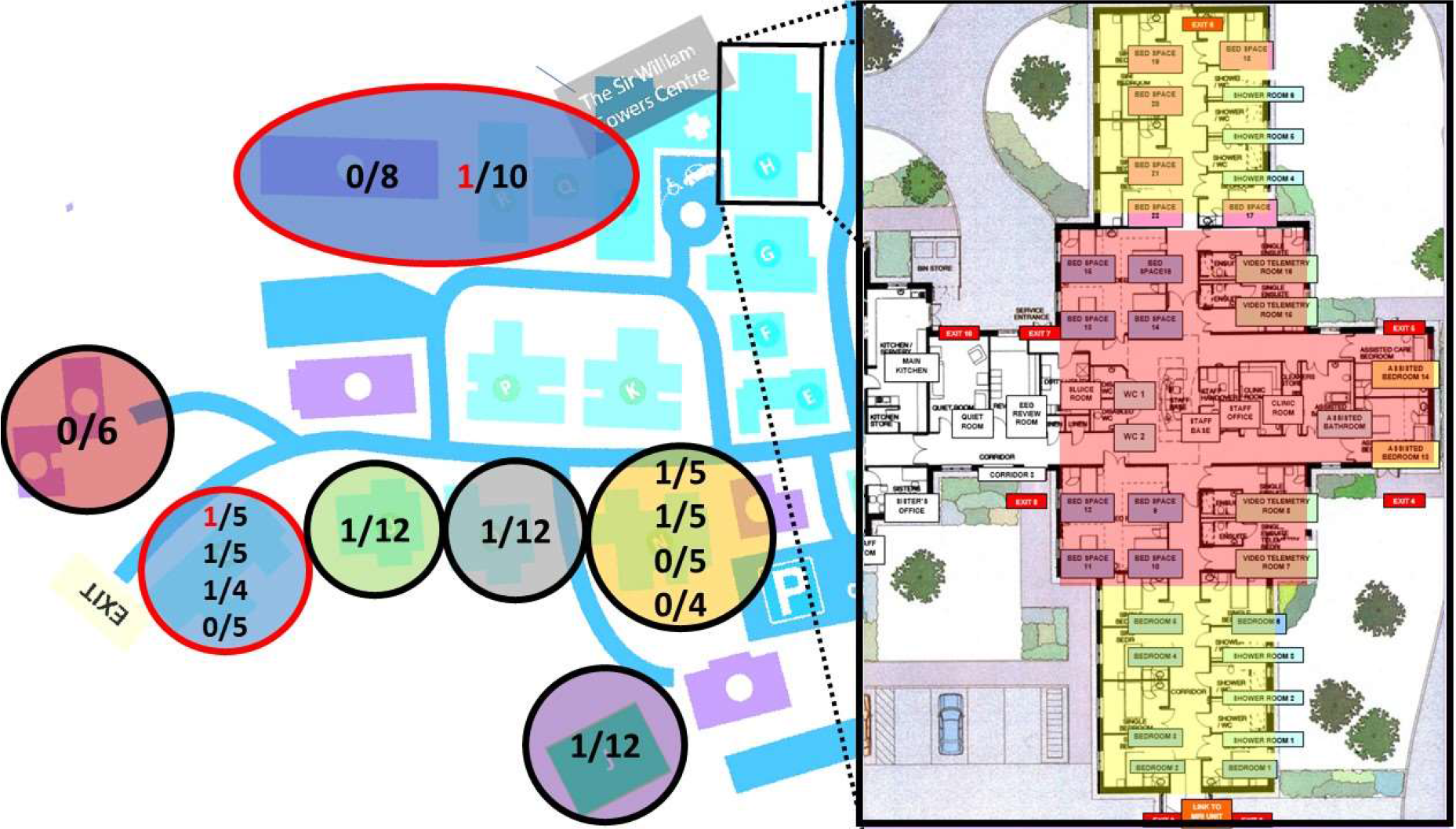
Map of Chalfont Centre for Epilepsy (CCE) site with enlarged illustration of the Sir William Gowers Centre (SWGC), the repurposed COVID-19 care unit at CCE. CCE houses 98 people who live in seven units of 1-4 self-contained flats. Outbreaks were observed in six of the seven units (represented as circles in different colours), with two of the nine positive residents that developed symptoms of COVID-19 (red numbers in red circles). Enlarged on the right of picture, Sir William Gowers Centre (SWGC), with six single rooms and eight beds ward repurposed for residents who tested positive (red area), and twelve beds for suspected cases who could not be isolated in their care homes (yellow).

St Elizabeth (STE), north-east of London, is a long-term care facility for 101 adults with severe epilepsy and other comorbidities. The adult residential facility consists of 11 units for 5–10 people, housing currently a total of 88 people (31 males), aged between 19–80 (median age: 42 years).

The Meath (TM), south-west of London, is a long-term care facility for 80 adults (median age 39 years, range: 23–79; 28 males) with epilepsy and additional learning and other disabilities. They live in nine residential units each housing between 3–13 people. UCLH provides tertiary care for 12/80 residents.

In response to COVID-19, different sets of measures were implemented on a short timescale (starting in mid-March) to keep those in the facilities as safe as possible, with limited resources. The measures fell into the categories of prevention and surveillance (Table 1), and intervention.

**Table 1.** List of prevention and surveillance measures adopted in the three care facilities starting on 23^rd^ March 2020. Chalfont Centre for Epilepsy (CCE), St. Elisabeth (STE), and The Meath (TM).

| **Prevention** |  |  |
| --- | --- | --- |
| Vulnerable people living in the facility-related | Staff-related | General measures |
| Houses / Bungalows treated as “family units” with free movement within that space (all centres), but encouragement of elderly individuals to spend most of the time in their rooms, in particular for meals (CCE) | “Staff rostering” with designation and isolation of flats within each care unit as stand-alone, with contacts between staff or individuals from different units reduced | Caregivers allocated to one individual for whole duration of shift, minimization of contact, with multiple tasks to be performed during same contact, e.g. dispensing medication and checking temperature (CCE) |
| Banning of family members from site, provision of laptops to maintain on-line contacts | No external visitors (all centres)<br>Temperature checks and PPE supplied for all essential workmen when entering care units (CCE) | Minimization of numbers of staff down to safe levels, with remote working where feasible, e.g. for administrative staff (all centres)<br>No Bank / agency staff or 1:1 (TM) |
| Closure of on-site communal areas (recreation hall, social, therapy and art centres) with cessation of group activities, but maintaining activities within the houses | PPE for all carers and other essential staff (e.g. cleaners) when entering all units (CCE)<br>PPE in use for personal care and administering emergency medications, and in isolation units at all times (STE, TM) | Social distancing for all activities as far as possible: staff required to keep 2 meters distance with other team members, except in special circumstances, e.g. an individual requiring support from more than one caregiver |
| Maintenance of activities with regular outdoor activities (closed to external visitors), e.g. walks in the gardens, listening to or playing music outside | Implementation of enhanced hygiene measures: regular cleansing of frequently touched surfaces, especially door handles | To wear aprons and gloves for close (<2 meter) contact with vulnerable individuals, with regular hand hygiene before and after,<sup>30</sup> eye protection where there is risk of contamination from respiratory droplets or from splashing of secretions (CCE) |
| **Surveillance** |  |  |
| Regular monitoring of body temperature (two/three times daily) of all those in care. Temperature > 37.8°C notified to the nursing and medical team for closer observation and escalation of isolation (see Figure 1 for CCE) and treatment | Regular monitoring of temperature of all caregivers and health care professionals at the start of each shift. No caregivers allowed to work if their temperature exceeded 37.5°C or if reported a new onset cough. Symptomatic carers immediately sent home to self-isolate for 14 days after symptom onset in line with PHE guidance. From April, at STE symptomatic carers and family members tested. |  |

### Intervention

At CCE, a program of systematic action was implemented for isolation and on-site testing for COVID-19 suspected cases. Individuals were suspected to have COVID-19 if they had a temperature > 37·8°C, or a temperature rise of 1·5°C above their long-term average, and/or new persistent cough or shortness of breath. SWGC was repurposed as an isolation facility. Any individual with suspected COVID-19 was admitted to SWGC (Figure 1, yellow area). Samples were obtained by nasopharyngeal and oropharyngeal swabs and tested at the Crick COVID-19 Consortium (CCC) by PCR for SARS-CoV-2.^12^ While waiting for the test results (up to 48 hours), individuals were cared for by dedicated and familiar caregivers in long shifts (i.e. 12 hours) to reduce staff contacts. Staff employed PPE and measures recommended for caring for confirmed COVID-19 cases.^13,14^ Cases testing positive were transferred to a separate section of SWGC (Figure 1, red area) for provision of the usual care and management, with additional vital signs monitoring using NEWS.^15^ If the result of the first testing in a symptomatic case was negative, a Second test was performed after 24–48 hours. If the second testing was negative, other causes for raised temperature or other symptoms were re-considered (unless already indicated). De-isolation of negative cases took place only after 48 hours following the resolution of the symptoms. After three weeks of intensive shielding and pragmatic surveillance of all people living in the facility, a further management step became available. This consisted of repeat enhanced surveillance of the remaining 97 of those in care, for early identification of positive cases in the asymptomatic phase.^16^ Weekly rounds of enhanced surveillance testing of all those in care have been undertaken since 17 April 2020. Naso- and oropharyngeal swabs were collected and tested as above.^12^ Results were available within 12–48 hours and prompted isolation of identified positive asymptomatic cases in SWGC as described above (Figure 1, red area). Tracing and testing of caregivers who had been in contact with those who had tested positive but were asymptomatic, was started within 12 hours of the original positive result. As a further preventative step, routine surveillance of all asymptomatic caregivers working on-site was commenced on 30 April 2020.

At STE and TM, early preventative measures were implemented to different degrees, but no on-site testing was available initially, with residents only tested when admitted to hospital. Residents were isolated within their rooms whilst presenting with COVID-19 like-symptoms, and/or transferred to dedicated units upon return from hospital, if COVID-19 was confirmed. Testing for caregivers with symptoms became available at testing stations since mid-April 2020, on-site testing for symptomatic residents since early May.

### Patient and Public Involvement

In mid-March 2020, families, residents (where having mental capacity), and care staff were all informed on the plans for infection control and containment at each centre. The objectives and outcome measures were developed and informed by the concerns of most families and care staff about infection spreading in the facilities, also given severe outcomes described in care homes worldwide. Some residents were shielded at their families’ home (CCE: 4; STE: 13, TM: 2). All families and care staff were supportive of the preventative measures, including visit restriction, hand hygiene and use of PPE, if available.

### Data sharing

Anonymised individual data will be made available upon reasonable request by bona fide researchers.

## Results

We report the outcomes in 1026 people living and working in three different long-term care-facilities, home for 286 residents with an age range of 19–91 years.

### Testing of residents with symptoms suggestive of COVID-19

#### CCE

Detailed demographic data for CCE are provided in table 2.

**Table 2.** Summary of demographic and clinical details of residents leaving at Chalfont Centre for Epilepsy (CCE).

|  | All<br>(n=98) | SARS-CoV-2 positive<br>(n=9) | SARS-CoV-2 negative<br>(n=89) |
| --- | --- | --- | --- |
| Male gender n, % | 66 (67%) | 8 (89%) | 58 (66%) |
| Age in years, mean (range) | 49 (23-91) | 52 (33-69) | 49 (23-91) |
| BAME | 5 (5%) | 2 (22%) | 3 (3%) |
| Fever (>37.8) and/or<br>respiratory symptoms n, % | 6 (6%) | 2 (22%) | 5 (5%) |
| Asymptomatic |  | 7 (78%) | 84 (95%) |
| Clinical frailty scale (1-9)<br>mean (range) | 5.88<br>(4-8) | 5.6<br>(4-8) | 5.91 (4-8) |
| Cardiac co-morbidity | 15 (15%) | 1 (11%) | 14 (15%) |
| Chronic respiratory<br>disease | 21 (21%) | 2 (22%) | 19 (22%) |
| Immunosuppression | 6 (6%) | 0 | 6 (7%) |
| Death | 1 (1%) | 1 | 0 |

Legend: BAME – Black, Asian and Minority Ethnic

By 10 April 2020, two COVID-19 symptomatic cases were identified amongst the 98 residents (2%) (Figure 1).

The first (#1–1) tested positive on 03 April and was an individual in their 60s, living in a large nursing home consisting of two units with 9–10 people each. This case had severe epilepsy and multiple comorbidities, including dysphagia with percutaneous endoscopic gastrostomy (PEG) in situ. They became symptomatic on the evening of 02 April, with vomiting and subsequently pyrexia at 39°C possibly related to aspiration, rapid and severe clinical deterioration with reduced oxygen saturation at ∼70%, persistent high temperature not responsive to paracetamol, reduced conscious level (Glasgow Coma Scale <5). Transfer to hospital was promptly arranged and the person tested positive on 03 April, following further deterioration, death occurred six days after symptom onset.

The second (#1–2) was an individual in their 60s, with a genetic epilepsy and comorbidities who lived in a large unit of 19 people with four self-contained flats each housing 4–5 people. On 09 April, they became pyrexial (38·7°C) and were promptly isolated in a single room in SWGC, tested and confirmed positive. They remained clinically stable until day 3, when oxygen saturation dropped to ∼85% leading to a transfer to our linked hospital facility (UCLH), given the risk of further deterioration. They tested positive again on days 7, 14 and 18, but remained clinically asymptomatic following admission, without pyrexia, and discharged back to CCE on day 40, after testing negative on two consecutive occasions.

The objectives and outcome measures were developed and informed by the concerns of most families and care staff about infection spreading in the facilities, also given severe outcomes described in care homes worldwide. Some residents were shielded at their families’ home (CCE: 4; STE: 13, TM: 2). All families and care staff were supportive of the preventative measures, including visit restriction, hand hygiene and use of PPE, if available.

As of 05 June, five other residents were promptly isolated due to the development of temperature above 37·°C, with or without respiratory symptoms: all have repeatedly (minimum twice) tested negative and were discharged back to their residences and de-isolated 48 hours after symptom resolution.

#### STE

By 7 May 2020, three symptomatic individuals were identified amongst the 146 people living on-site (2%).

The first (#2–1) was a young adult with epilepsy following encephalitis aged 2, dysphagia with PEG in situ and severe LD who lived in a unit with eight other people. He was admitted to hospital on 05 March, with aspiration pneumonia following an episode of vomiting, tested then negative, and was discharged 09 March. Two weeks later, on 23 March, he presented with a new cough and pyrexia (37·9°C), was transferred back to the hospital the same day, and then tested positive. He required ventilation. Death occurred 11 days after symptom onset

The second (#2–2) was an individual in their 50s, with a genetic epilepsy who lived in the same unit as #3. On 09 April, this individual became symptomatic with fever (39·0°C), lethargy and cough for 1 week, after which they rapidly deteriorated with respiration rate > 32 and oxygen saturation <88%. He was promptly isolated and confirmed positive on 20 April. He remained in isolation until 05 May. One carer at the same unit showed symptoms on the same day as #2–2, and tested positive. Another carer was asymptomatic and tested positive on 08 May.

The third (#2–3) was an individual in their late 50s, with refractory epilepsy of unknown cause, moderate LD who lived in a different unit to #2–1 and #2–2. The individual became symptomatic on 22 April, with mild fever (37·8°C) and cough, but would not consent to isolation in room and so was moved to an unused area of another building. Supplemental oxygen was used for the first few days as his oxygen saturation fell <90%, but, overall, symptoms remained mild. A positive result for COVID-19 testing was received on 01 May.

As of 05 June, eight other individuals were promptly isolated due to the development of temperature above 37·8°C, with or without respiratory symptoms, only six of those eight were tested once, all negative. All eight individuals were discharged back to their residences and de-isolated 24–48 hours after symptom resolution.

#### TM

By 05 June 2020, eight symptomatic individuals were identified amongst the 80 people living on-site (10%). Symptoms included fever above 37.8°C, cough, and other respiratory symptoms. There was no access to viral testing, but they were promptly isolated for at least 48 hours after complete resolution of the symptoms

### Testing of asymptomatic residents

On 17 April 2020, CCE started regular weekly surveillance of all individuals living onsite. Of the remaining 96 people, seven were not tested in the first round: five declined, two had temporarily moved back to live with their families. Of the 89 tested, four were found to be positive (4.5%), and were immediately isolated.

On 22 April, in the second surveillance round, we tested 95/96 people; only one continued to decline testing. An additional three asymptomatic individuals tested positive who had previously tested negative five days earlier. All three remained asymptomatic for COVID-19 as of 05 June.

On 27 April, in the third surveillance round, all 96 people tested negative, including one of the seven asymptomatic residents who had tested twice positive before. All seven cases have since tested negative twice 24–48 hours apart.

Since 05 May, during a further five rounds of weekly surveillance, we tested all 96 (97 since 26 May) people, and all were negative on each occasion.

At STE, a fourth (#2–4) individual tested positive on 07 May, during one of their frequent hospital admissions for recurrent urinary tract infections, but was considered asymptomatic for COVID-19 as malaise was attributed to the other health conditions, and was tested negative prior to discharge on 13 May. This individual in their late 40s lives in a different unit than the three symptomatic residents tested positive. One carer from the same unit became symptomatic on 11 April and another on 08 May, both tested positive.

There was no routine asymptomatic screening at STE or TM during the observation period.

### Contact tracing and surveillance of care staff

At CCE, following confirmation of positive results, contact testing of all cares who had been in contact over the previous two weeks with the individuals who tested positive was performed within three days. In total, 150 caregivers accepted testing, only one tested positive. From 30 April onwards, weekly surveillance of all asymptomatic 275 caregivers has been implemented: none has been positive on any occasion.

At STE, from 06 April onwards, testing was available for symptomatic carers and those needing to self-isolate for 14 days because a member of their household had symptoms. Contact tracing was implemented from 02 May, with testing of all carers who had contact with the residents who have been tested positive. Out of the 215 staff, 105 people were tested once, 14 positive symptomatic or self-isolating caregivers were identified, with an additional asymptomatic carer found positive after introducing contact tracing.

At TM, up until 05 June, 26 of 250 staff were symptomatic, and have been tested with two positive results.

## Discussion

We report confirmed COVID-19 outbreaks in two out of three care facilities for people with epilepsy and additional co-morbidities with only 2% of residents (CCE: 2/98; STE: 3/146) showing COVID-19 related symptoms and testing positive. Enhanced surveillance, available at CCE, revealed a high rate of asymptomatic SARS-CoV-2 infected residents (7/9 tested positive; 78%). Our case fatality rate was high (CCE: 50%, or 11% corrected for asymptomatic; STE: 33%), but total number of deaths, one at each of the two centres, was in line with the average death rate for a 12 weeks observation period over the last five years.

Our observations at CCE of a relatively low (9%) infection but high (78%) asymptomatic rates are similar to the report of initially heathy populations (3711 passengers on Diamond Princess cruise ship) with 19% testing positive and of those 47% being asymptomatic.^18^ Our higher asymptomatic rates might be explained by the difficulties of detecting mild or no symptoms in people with severe LD. However, our rates are very different from those reported in another, similar sized long-term care facility with access to testing asymptomatic residents: among 76 residents, 48 (63%) tested positive initially with 27 (56%) asymptomatic at time of testing, but only three remained asymptomatic (6%).^4^ Their case fatality rate was also higher (26%), possibly due to a difference in population characteristics (average age of those tested positive: 79 years versus 52 years at CCE).

We succeeded in containing a wide-spread outbreak of SARS-CoV-2 in six of seven care units at CCE with a low rate of spread, i.e. only one infected resident per individual care unit, with no established resident-to-carer transmission. Only one carer tested positive during immediate contact tracing and none of the 275 carers since weekly surveillance was implemented, suggesting that the widespread outbreak was effectively contained within three weeks. In contrast, at STE without enhanced surveillance, 15 tested positive out of 105 symptomatic carers tested once since testing of symptomatic carers became available at STE on 06 April. Infections of residents and carers were widespread across almost all care units at both, CCE (6/7) and STE (11/11). However, whilst the spread of infections was contained at CCE within 3 weeks, positive test results at STE were encountered throughout the 12 weeks’ observation period (see figure 2). Whilst symptom severity was similar between the two sites, we conclude that the difference in numbers of infected staff is likely due to enhanced surveillance available at CCE, which allowed identification, and consecutively isolation of asymptomatic residents. An alternative explanation would be that asymptomatic people are less likely to transmit, but this would not explain the difference in numbers of symptomatic staff between CCE and STE.

**Figure 2.**
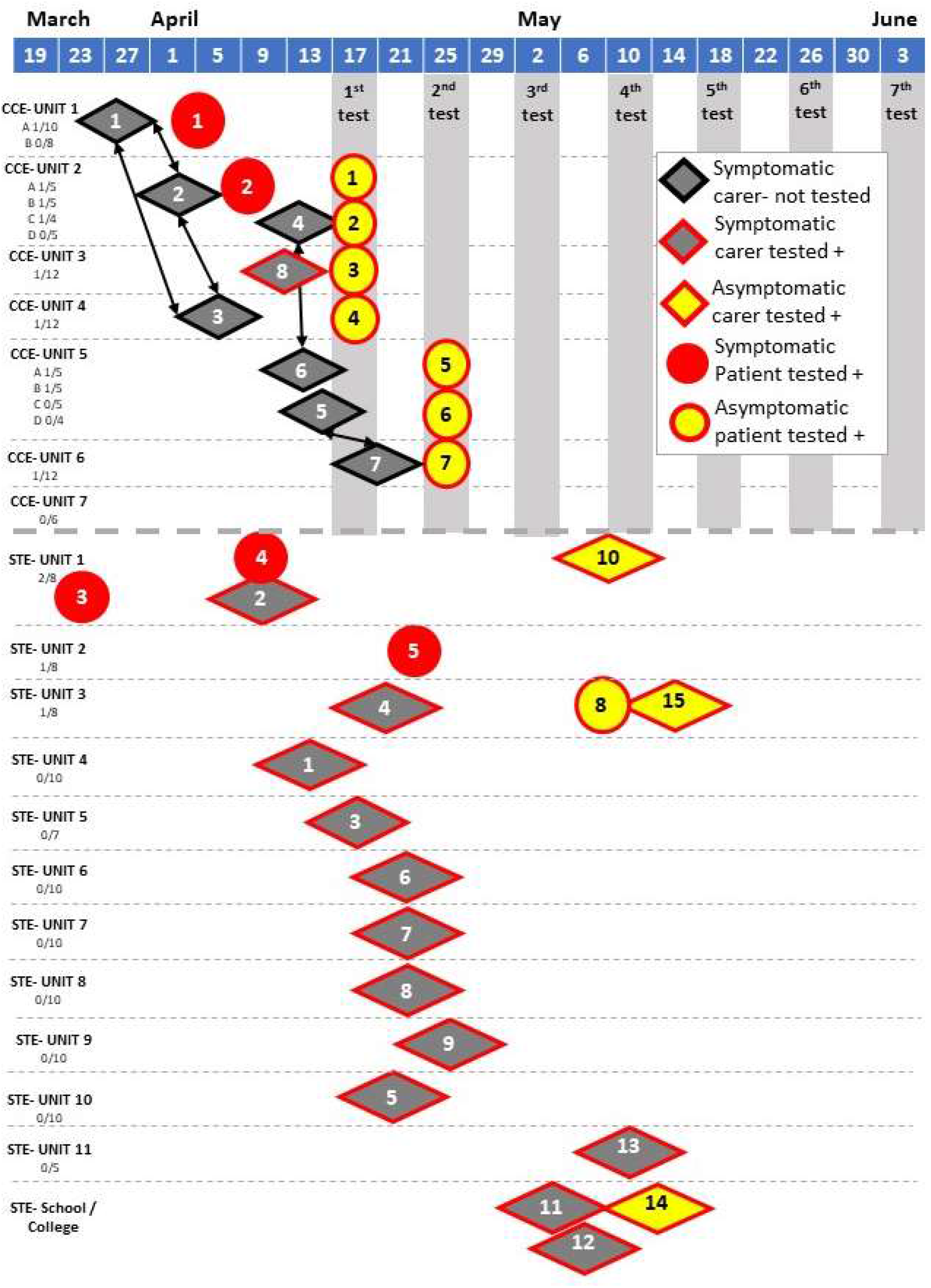
Timeline across centres CCE and STE. This includes all symptomatic cases tested positive (red circle 1–5), asymptomatic tested positive (yellow circle 1–8), selected symptomatic staff at CCE (grey diamond 1–7, self-isolating but not tested), and symptomatic staff at STE tested positive (red outlined grey diamond 1–9,11–13) and asymptomatic staff (red outlined yellow diamond 10,14,15). Staff are presented in the unit where they regularly worked, arrows connect staff who are also household contacts at CCE. Timings represent date of symptom onset (symptomatic patients), or date of self-isolation from work (staff members, who were not PCR tested), grey columns represent date of asymptomatic case screening,

Care facilities are highly vulnerable to COVID-19 outbreaks,^9,10,19^ and it is crucial to identify effective strategies to prevent infection and to reduce impact. The approach reported here focused on two main strategies: (1) early on-site enhancement of preventative and infection control measures, (2) early identification and isolation of symptomatic cases, with enhanced surveillance and isolation of asymptomatic people living and working at CCE as an additional measure. All centres were able to implement isolation of suspected and confirmed cases and to enforce the use of PPE through open market sourcing rather than waiting for centralized procurement.^20^ Similar early implementation of these measures in a care-facility in the US has been reported to be effective in minimizing viral spread.^21^ Whilst this is reassuring, suggesting that PPE and good hand hygiene have been used effectively when in contact with confirmed positive individuals, carers themselves must have been pre- or asymptomatic earlier and so, unknowingly, infected colleagues and residents under their care, as it happened at CCE. The initial spread of infection across the sites, very likely caused by healthcare workers from different care units sharing accommodation (see figure 2), questions the initial advice to healthcare workers of continuing to go to work despite household members self-isolating.

Residents in all centres had different degrees of LD, such that it was not possible to assess reliably for the presence of non-respiratory symptoms, which have been described involving various organs.^3,22^ For example, acute-onset anosmia may manifest either early in the disease process or in people with mild or no constitutional symptoms.^23^ Thus, enhanced surveillance through repeat testing of all ‘asymptomatic’ cases is vital for case ascertainment in such settings, but can be unreliable at times, even if symptomatic, due to the degree of LD. Similarly, due to limited compliance the false negative rate of testing can be expected to be higher in this population than the already quoted 20–30%. Thus, repeat testing is crucial, three of the seven asymptomatic SARS-CoV-2 positive residents at CCE tested negative during the first round of surveillance.

Screening of asymptomatic cases and testing after symptom resolution is also crucial to identify covert transmitters and individuals at risk of rapid deterioration.^24^ According to UK public health guidance, a negative test was not required prior to discharge from hospital back to a care-facility.^26^ Such discharges may contribute to the risk of infection spreading within care-facilities.

Challenges were encountered when setting up a strategy for a group of people with significant LD, autism or challenging behaviour, which complicate attempts to isolate. Social care staff in residential homes were not necessarily familiar with barrier nursing or infection control, and only a minority had nursing training. Ongoing surveillance of symptoms with regular temperature monitoring, and early isolation and testing of symptomatic people suspected of COVID-19 is key here to infection control, but required changes in organisational structures within units. Mitigations of these difficulties included the proactive re-purposing of SWGC (CCE) or empty units (STE, TM) as isolation facilities for those for whom this would have not been possible in their residential units. Continuity of care by staff acquainted with the individuals was deemed essential for the residents’ wellbeing, and for the proper evaluation of symptoms and presentations.

Despite the frailty and multiple co-morbidities of our population, the impact of SARSCoV-2 in all the facilities has been limited to date. Children and young adults appear to have lower infection rates, although access to testing, even of symptomatic residents, was limited in this age group. Enhanced surveillance, like at CCE, is required to determine the true infection rate in the younger age groups. Three of the confirmed positive at CCE/STE have an underlying genetic condition frequently observed in children with severe epilepsy, with mutation in the SCN1A gene, which is known to be associated with fever sensitivity and elevated risk of early mortality.^27^ Host genetic predictors of outcome in SARS-CoV-2 infections are yet to be established.^28^ SARS-CoV-2 RNA mutations and additional molecular mechanisms may explain variability in clinical presentation.^28–30^

Not surprisingly, contact tracing at CCE proved difficult, not only for asymptomatic residents testing positive without data on when the infection might have occurred, but also due to delay in obtaining test results (up to 5 days after testing), carers sharing accommodation (contacts of contacts, see figure 2) and large numbers of agency workers, in particular in CCE-Unit 2. Testing of symptomatic carers at STE (12 positive out of 105 tested) and TM (2/26) returned similar numbers of positive tests in symptomatic people compared to the general UK population (as of 04 June 2020: 284,868 cases / 5,438,712 tests), with the official numbers not accounting for multiple tests in hospitals for the same patient (two negative tests prior to discharge). Together with a low rate of infected residents, this is reassuring as it suggests that early implementation of preventative and infection control measures in all three long-term care-facilities (see table 1) can reduce the infection risk in high-risk environments^11^, be it for vulnerable individuals living in long-term care facilities or their carers, to a level similar to that observed in the general population. However, we also show that these measures alone, without identification of asymptomatic people through enhanced surveillance, do not contain the spread of infection.

Here, we provide the evidence of the need for enhanced surveillance for SARS-CoV-2 of asymptomatic persons in high risk environments. We recognize that CCE was fortunate to have extensive collaboration between basic science repurposed for high-throughput viral testing (the Francis Crick Institute), high-level virological and clinical input (from UCLH), and the ability to redeploy clinical academics (from UCL), to support dynamic and purposeful care teams. All centres benefit from close integration between health and social care with close reviews by epilepsy consultants from UCLH and/or GOSH. Such multidisciplinary input is not available to all care facilities, but the strategies outlined here may provide generally applicable guidance to other facilities facing similar challenges, in particular in preparation for a potential second wave of infection. We hope that such integration between science, healthcare and social care can also generate a new model for the care of the most vulnerable in society in the future. We must learn that there are better ways to be a civil society, to ensure that those living in care-facilities are not excluded from the expertise and interventions available for the wider population.

## Data Availability

Anonymised individual data will be made available upon reasonable request by bona fide researchers subject to NHS requirements.

## Acknowledgements

We thank all care staff working, and all people living at the Chalfont Centre for Epilepsy, St Elisabeth, and The Meath, and their families. S Balestrini, MJ Koepp, JW Sander and S Sisodiya are based at UCLH/UCL Comprehensive Biomedical Research Centre, which receives a proportion of funding from the UK Department of Health’s NIHR Biomedical Research Centres funding scheme. JW Sander receives support from the Dr Marvin Weil Epilepsy Research Fund, the Christelijke Verenigingvoor de verpleging van Lijdersaan Epilepsie, The Netherlands, and the UK Epilepsy Society.

